# Understanding Cardiac Events in Breast Cancer (UCARE) - Pilot Cardio-Oncology Assessment and Surveillance Pathway for Breast Cancer Patients

**DOI:** 10.1101/2023.12.15.23300055

**Authors:** Michael Cronin, Aoife Lowery, Veronica McInerney, William Wijns, Michael Kerin, Maccon Keane, Silvie Blazkova, Michael Martin, Osama Soliman

## Abstract

In Ireland, over 3,000 patients are diagnosed with breast cancer annually, and 1 in 9 Irish women will be diagnosed with breast cancer in their lifetime. There is evidence that female breast cancer survivors are more likely to die of cardiovascular disease than their age-matched counterparts. This protocol describes a prospective, single arm, pilot feasibility study implementing a dedicated Cardio-Oncology assessment and surveillance pathway for patients receiving multimodal breast cancer treatment. It incorporates novel biomarker and radiomic surveillance and monitoring approaches for cancer-therapy related cardiac dysfunction into routine care for breast cancer patients undergoing adjuvant systemic chemotherapy.

## Introduction

Breast cancer is the commonest female malignancy in Europe (1). Each year more than 2 million women are diagnosed with breast cancer globally with about 685,000 deaths from the disease in 2020 (2). In Ireland, over 3,000 patients are diagnosed annually, and 1 in 9 Irish women will be diagnosed with breast cancer in their lifetime (3). Multimodal oncological and surgical practice has improved outcomes, making breast cancer highly curable if diagnosed and treated appropriately at an early stage (4). Data from the National Cancer Registry confirms a decline in age-standardised mortality rates for breast cancer and a marked improvement in 5 and 10-year survival rates over the past 2 decades (3). These improvements in breast cancer outcomes are due to earlier diagnosis with the introduction of screening programmes and therapeutic advances informed by an improved understanding of breast cancer molecular biology (3, 5).

As breast cancer specific mortality improves, there is an increasing recognition of both other causes of mortality and unmet survivorship needs in patients living with and after breast cancer treatment. This is particularly evident in patients for whom the success of breast cancer treatment is tempered by the sequelae of cancer treatment received. There is evidence that female breast cancer survivors are more likely to die of cardiovascular disease than their age-matched counterparts (6). This relates not only to a higher prevalence of risk factors for cardiovascular disease such as dyslipidaemia, abdominal adiposity, hypertension, and diabetes mellitus in the breast cancer population (7), but is increased exponentially by Cancer Treatment-Related Cardiac Dysfunction (CTRCD), particularly in patients with Her2 positive breast cancer exposed to multimodal cancer therapy.

CTRCD refers to the cardiotoxic effects of cancer treatment which can occur acutely during cancer treatment, disrupting the treatment course, or as a late effect which can adversely affect quality of life and overall survival (8). The combination of cytotoxic chemotherapy, radiotherapy and Anti-HER2 therapy (Trastuzumab) administered to patients with HER2 -positive breast cancer poses a significant cardiotoxic risk, particularly of left ventricular dysfunction and heart failure (8). These risks are even higher in an ageing breast cancer population in whom pre-existing HF is increasingly common (9) and several large prospective analyses describe older patients performing worse than their younger counterparts, with cardiopulmonary disease contributing to poorer outcomes (10, 11).

Management of these patients requires a careful balance of adequate and appropriate cancer treatment while minimising cardiac risk and there is evidence that early diagnosis and treatment of CTRCD increases the potential for minimising or reversing cardiotoxic effects (12). This emerging area of medicine has limited prospective evidence on which to base decision-making, however the European Society of Cardiology (ESC) have recently published guidelines which aim to assist healthcare professionals providing care to oncology patients before, during and after cancer treatment with respect to their cardiovascular health and wellness (13)

The Irish National Cancer Strategy 2017 – 2026 (14) recommends that models of care be developed to ensure that patients receive the required care from an expert multidisciplinary clinical team. Furthermore, the 2019 Irish National Cancer Survivorship Needs Assessment identified the impact of treatment side-effects as an unmet research need (15). Indeed the European Society of Cardiology state in the 2022 Cardio-Oncology guidelines (16) list within gaps in the evidence “Robust evidence on the impact of dedicated cardio-oncology programmes”. The Understanding CARdiac Events in Breast Cancer (UCARE) study addresses these needs by developing and pilot testing a structured Assessment and Monitoring Care Pathway at Galway University Hospital for patients at risk of developing cardiac toxicity from cancer treatment.

### Study Rationale

This prospective multi-centre randomized control trial is being done to find out if it is feasible to run a Cardio-Oncology Assessment and surveillance clinic in Galway for women with a recent diagnosis of breast cancer who will be receiving systemic chemotherapy as part of their treatment. The investigators will explore if a dedicated care pathway incorporating a comprehensive assessment of cardiac health before and during chemotherapy treatment will enable identification of those at higher risk of developing CTRCD. Cardiovascular complications from cancer therapies represent an important challenge for many cancer patients, both at the time of treatment as well as survivorship after treatment. Unfortunately, the investigation and management of cardiac disease in cancer patients is currently suboptimal. It is envisaged that early identification of high risk patients will allow protective strategies and intensive surveillance to be put in place to reduce risk and improve outcomes for these patients. This study has been assigned the ID NCT05921279 with clinicaltrials.gov.

The anticipated benefits from this research are:

- This pilot feasibility study will inform specific uncertainties including practicality of delivering the intervention (Cardio-Oncology assessment and surveillance pathway) in the existing clinical setting i.e. identifying facilitators and barriers for the intervention.
- Participation in this research will provide patients with more education on their cardiac risk during cancer treatment than if they did not participate.
- Patients will be risk stratified for CTRCD at baseline which may trigger referral to cardiac specialists or increased surveillance and earlier detection of CTRCD.
- Service development and service enhancement for cancer survivors: It is expected that the projects’ output will include evidence to support the value of establishing dedicated cardio-oncology clinics and the significant role that advanced nurse practitioners/allied health professionals can play in cancer survivorship. Output will include the development of a cardio-oncology nurse specialist role.
- Educational program development: We will incorporate our results in clinical and academic programs. The development of a Cardio-oncology fellowship programme at Galway University Hospital, training the next generation of cardiologists to ensure that there is specialist expertise for the cardiovascular care of cancer patients.

### Study Objectives

#### Primary Objective

1. To evaluate the feasibility of implementing the intervention of a dedicated Cardio-Oncology assessment and surveillance pathway incorporating novel biomarkers, radiomic surveillance, and monitoring approaches for CTRCD into routine care for breast cancer patients undergoing adjuvant systemic chemotherapy.

#### Secondary Objectives

1. To evaluate the baseline cardiovascular health of patients undergoing multimodal treatment for breast cancer, including systemic chemotherapy.
2. To identify risk factors for the development of acute CTRCD.
3. To describe the incidence of acute CTRCD in patients receiving systemic breast cancer treatment.
4. To elucidate the role of specific imaging, clinical and laboratory markers in the prediction and early detection of CTRCD in patients.
5. To collect and biobank relevant specimens and data for biomarker discovery
6. To evaluate the baseline physical activity and quality of life (QOL) in patients referred for systemic chemotherapy and the QOL implications of CTRCD.
7. To assess mental health including stress/anxiety via wearable devices in patients with high risk for CTRCT receiving systemic chemotherapy.
8. To assess patient satisfaction of the Cardio-oncology pathway and to determine their perceptions of the pathway’s usability.

Outcome measures are available in following tables 1 and 2.

**Table 1.**
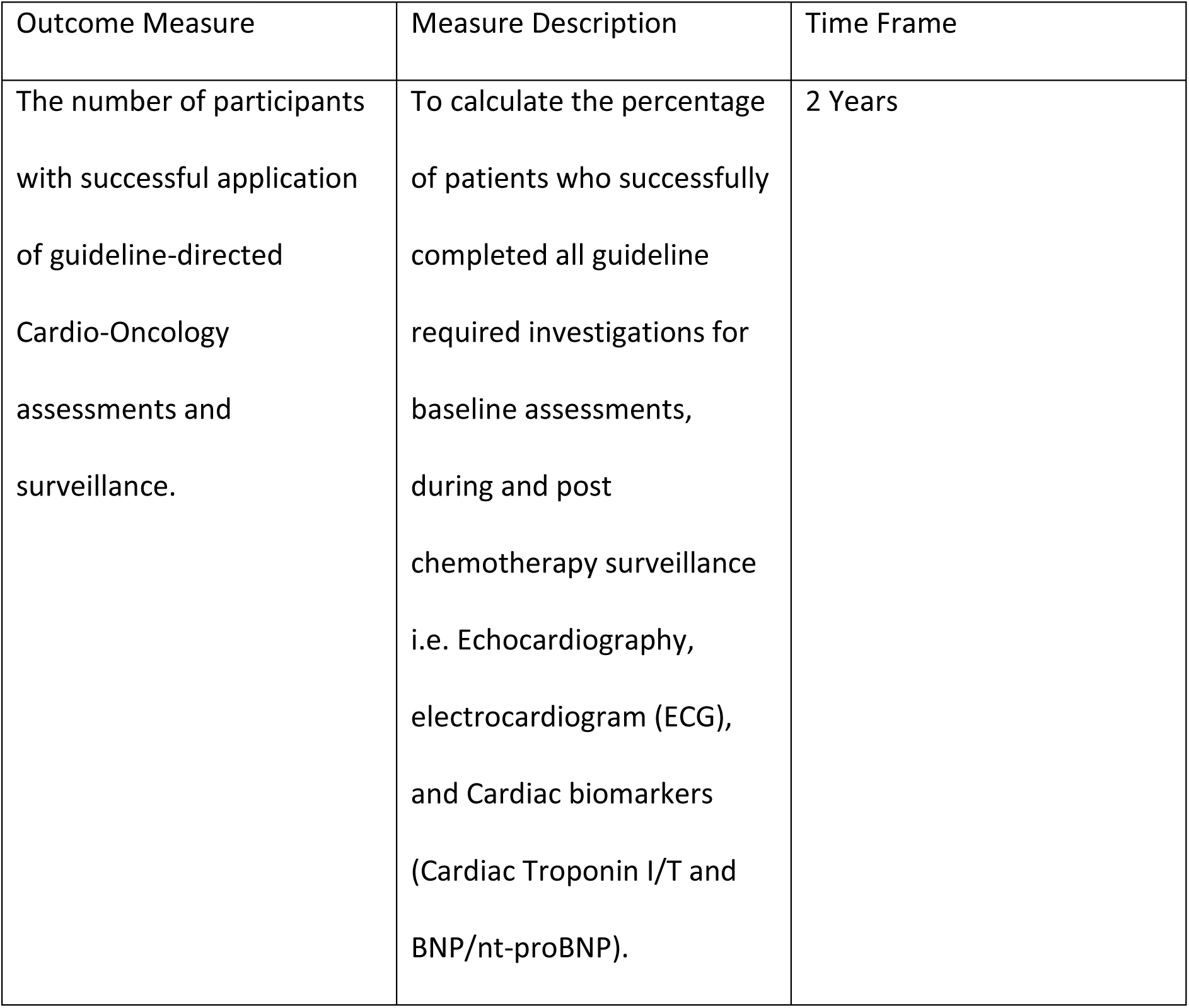
Primary Outcome Measures.

| Outcome Measure | Measure Description | Time Frame |
| --- | --- | --- |
| The number of participants with successful application of guideline-directed Cardio-Oncology assessments and surveillance. | To calculate the percentage of patients who successfully completed all guideline required investigations for baseline assessments, during and post chemotherapy surveillance i.e. Echocardiography, electrocardiogram (ECG), and Cardiac biomarkers (Cardiac Troponin I/T and BNP/nt-proBNP). | 2 Years |

**Table 2.**
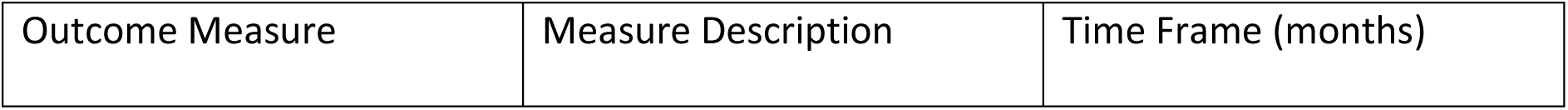
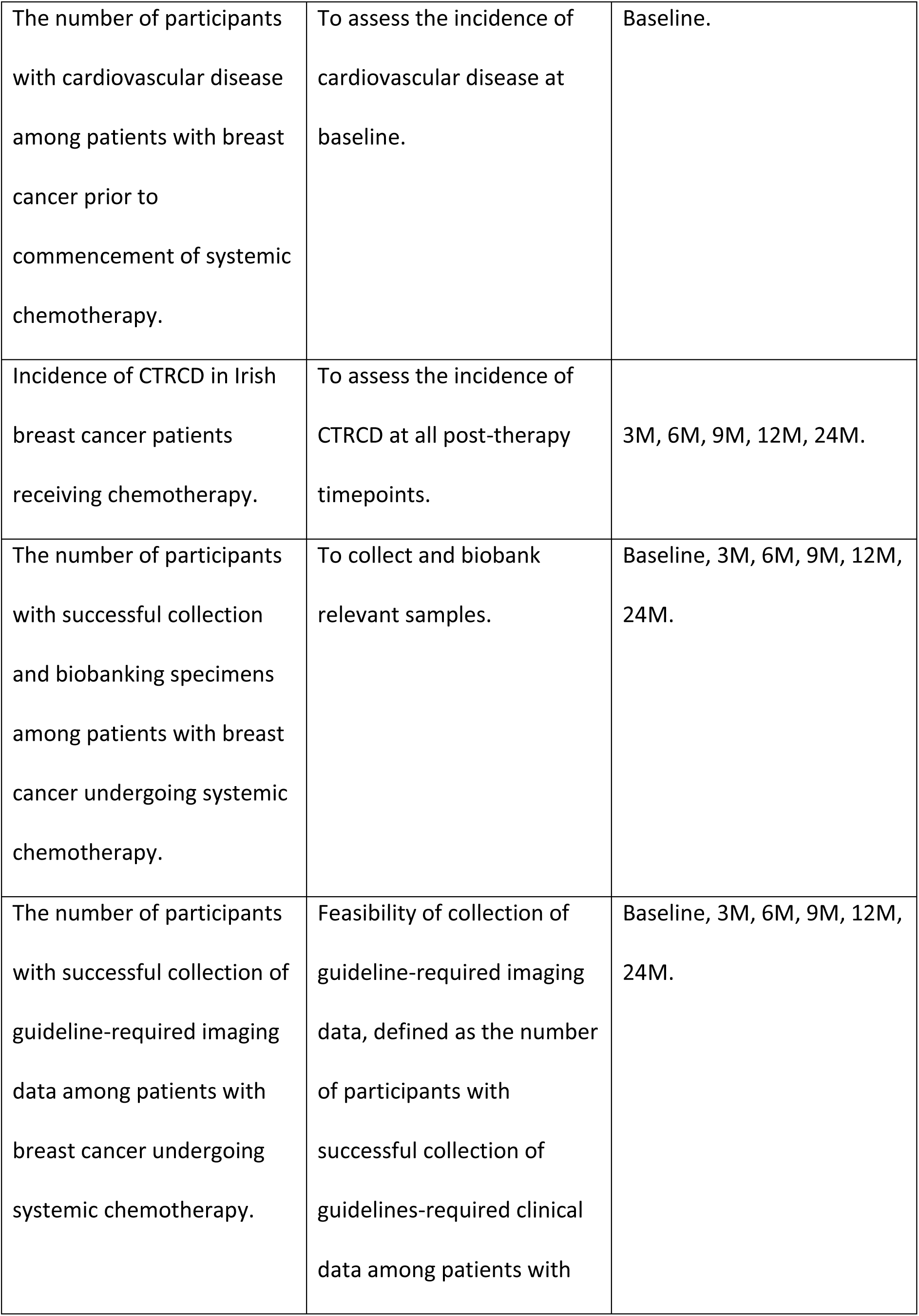
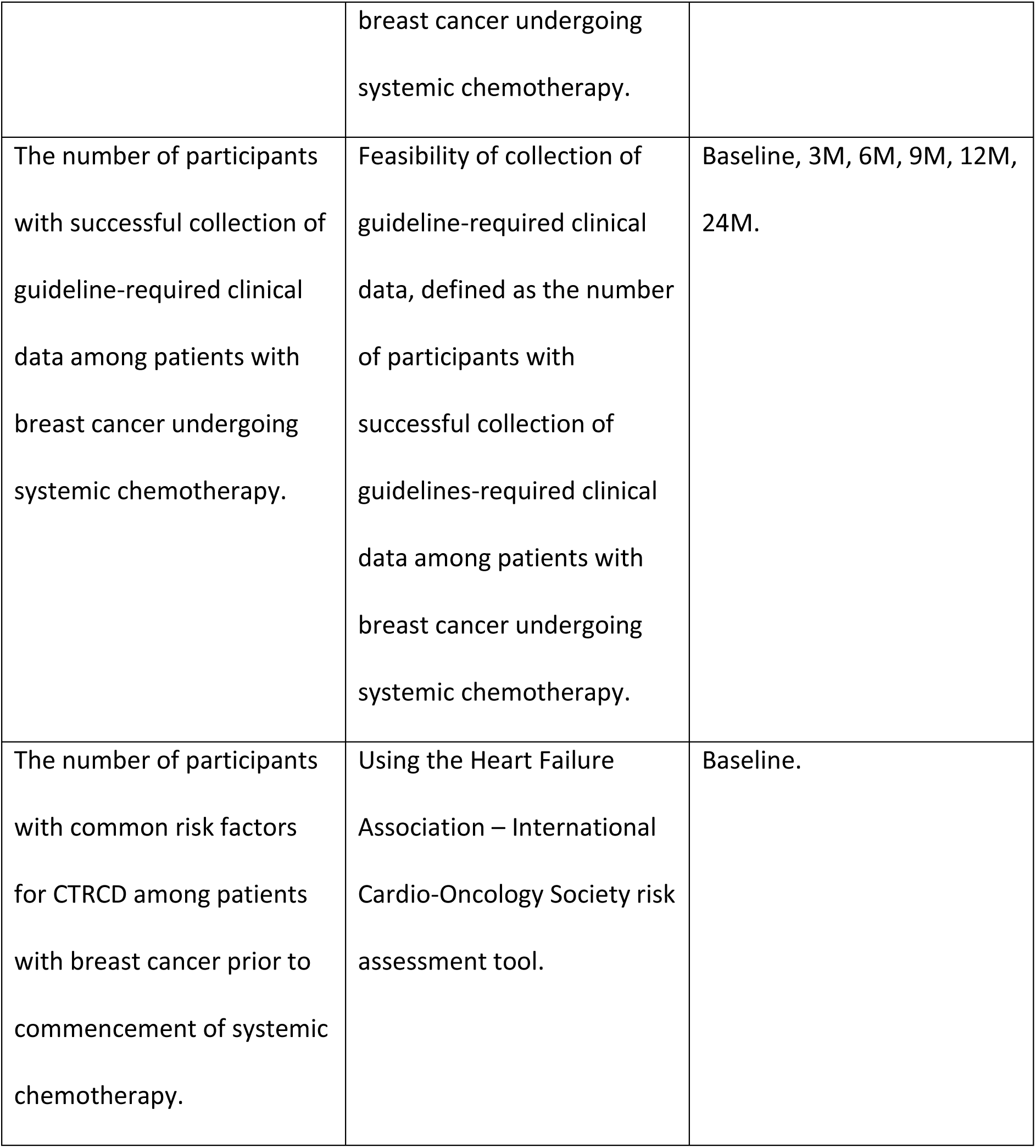
Secondary Outcome Measures.

## Methods

This study is a prospective, single arm, pilot feasibility study. The study will focus on adult female patients diagnosed with stage I-III breast cancer. It will take place over multiple sites: Galway University Hospital, Sligo University Hospital, and Mayo University Hospital, led by local physicians with the aid of dedicated research nurse specialists and advanced nurse practitioners. Weekly meetings will be held by the research team.

### Enrolment

During the initial 12 months of the study, adult female patients diagnosed with stage I-III breast cancer and referred for systemic chemotherapy will be enrolled at the time of their clinic visit for discussion and scheduling of systemic therapy. Patients in residential care or patients with life-threatening health conditions will be considered for inclusion, however if there is an acute life-limiting condition threatening the patient’s life at the time of consent they will not be considered for inclusion in the study.

Inclusion Criteria:

- Women aged ≥ 18 years.
- Ability to read and understand English.
- Breast Cancer Stage I - III planned to receive systemic chemotherapy.

Exclusion Criteria:

- Patients who are not for systemic chemotherapy with curative intent.
- Patients who are unable to co-operate with the study protocol.
- Patients who are unable to give informed consent.

### Consent

Patients will be consented by a member of the study team (cardio-oncology research nurse). The patient information leaflet (PIL) and informed consent form (ICF) can be found within the supplemental material. This may occur in person, as they present for their clinic visit.

Alternatively, this may involve a remote consenting process (e.g pending geographic or illness-related barriers) as follows:

- The potential participant will receive a hard copy of the PIL/ICF by email or post.
- A member of the study team will telephone the potential participant (or may use online communication platform), going through the PIL and answering any questions which participant may have.
- If the patient is happy to take part the researcher will ask them to sign and date the ICF and send it back to the study team by email or by post.
- The member of the study who explained the study to the patient will then sign and date the ICF.
- The completed ICF (now signed and dated by both participant and research team member) will be filed in the study file which is kept securely. One copy will be placed in the patients’ medical notes and one copy will be emailed or posted back to the participant for their own records.

All enrolled patients will receive a cardiovascular assessment at baseline as per the 2022 ESC guidelines (16). There are 4 study group cohorts amongst the study (see table 3). Each cohort will be monitored by collecting biobank bloods, ECG and transthoracic echocardiography (TTE) as per the ESC guidelines whilst receiving chemotherapy. The specific protocol for each group can be found in the supplementary material. Risk stratification will be via the Heart Failure Association – International Cardio-Oncology Society (HFA-ICOS) cardiac risk assessment tool (17). If consented, correspondence will be sent to the patients General Practitioner with a brief description of the study outline (see supplemental material). Correspondence will be sent to the primary medical oncologist treating the patient, with a description of their risk category and pathway selection (see supplemental material).

**Table 3.** Cohorts.

| Group | Cancer Therapy | Risk |
| --- | --- | --- |
| 1 | Anthracycline based chemotherapy | Low and moderate risk |
| 2 | Anthracycline based chemotherapy | High and very high risk |
| 3 | Herceptin targeted therapy | Low and moderate risk |
| 4 | Herceptin targeted therapy | High and very high risk |

### Cardiac Risk Assessment and Surveillance

Surveillance protocols will be led by the ESC 2022 guidelines in cardio-oncology across the 4 included groups (see supplementary materials). All patients will receive a comprehensive physical examination and history at follow up visits. Cardiopulmonary exercise testing will be undertaken in patients with a clinical indication. Otherwise physical activity will be assessed and monitored using the international physical activity questionnaire (see supplementary material). The health of the participants will be monitored as per standard treatment guidelines by the attending medical oncology, cardio-oncology, and surgical oncology clinical teams. These teams will be responsible for their patients as typically occurs in standard clinical practice. Patients who in whom CTRCD is identified during cardiac surveillance on this pathway will be referred and managed appropriately as per ESC guidelines and current best practice.

Regarding long term follow-up after the study is completed, the health of the participants will be monitored as per current best practice guidelines by the attending medical oncology follow up clinics as per standard practice. Those patients who require dedicated cardiac follow up will be referred accordingly as per best practice ESC guidelines.

### ECG

Baseline 12 lead ECG will be undertaken for all patients, to include measurement of QT interval (corrected). This will be undertaken by the cardio-oncology research nurse at the baseline assessment. An abnormal baseline ECG will trigger referral to a cardiologist. A Stress ECG will also be undertaken in patients when clinically indicated.

### Questionnaires

Patients will be emailed a link to the EORTC QLQ-BR45, QLQ-30, IPAQ-SF, KCCQ-12, PSS-10 and STAI questionnaires at 3 monthly intervals over the course of 24-month period. These surveys will be accessible through any standard internet browser. Patient Reported Quality of Life Outcomes will be collected using an electronic data capture system (REDCap). This will be facilitated through the Clinical Research Facility in Galway University Hospital, meeting national and international guidelines in relation to data privacy and protection standards.

Additionally, patients, healthcare professionals and the team involved in the development and implementation of the pathway will be invited to attend either a focus group, or one to one interview, that will aim to assess experiences of the cardio-oncology care pathway, associated processes, limitations, challenges and possible benefits or recommended changes to the pathway. The patient-reported outcomes will include the following (which can be found in the supplementary material):

#### 1 The European Organisation for Research and Treatment of Cancer (EORTC) QLQ-C30 and QLQ BR 45

The QLQ-C30 comprises 30 items that can be summarized in 15 scales: physical functioning, role functioning, social functioning, emotional functioning, cognitive functioning, global QOL, fatigue, pain, nausea/vomiting, appetite loss, dyspnoea, sleep disturbances, diarrhoea, constipation, and financial impact of disease. Overall the design is to measure to assess QOL among cancer patients (18).

Disease-specific details will be collected using EORTC modules for breast i.e. QLQ BR 45.. This questionnaire incorporates nine multi-item scales to assess body image, sexual functioning, breast satisfaction, systemic therapy side effects, arm symptoms, breast symptoms, endocrine therapy symptoms, endocrine sexual symptoms. In addition, single items assess sexual enjoyment, future perspective and being upset by hair loss.

#### 2 International Physical Activity Questionnaire Short Form (IPAQ-SF)

IPAQ-SF assesses physical activity undertaken across a comprehensive set of domains. The specific types of activity that are assessed are walking, moderate-intensity activities and vigorous intensity activities. The frequency (measured in days per week) and duration (time per day) are collected separately for each specific type of activity. This measure assesses the types of intensity of physical activity and sitting time that people do as part of their daily lives, and can be considered to estimate total physical activity in METS per time unit, and lastly time spent sitting

#### 3 Kansas City Cardiomyopathy Questionnaire (KCCQ-12)

KCCQ was designed with input from patients and clinicians to capture those domains of how heart failure affects patients’ lives. The original KCCQ includes 23 items that map to 7 domains: symptom frequency; symptom burden; symptom stability; physical limitations; social limitations; quality of life; and self-efficacy (the patient’s understanding of how to manage their heart failure). To make the KCCQ more feasible to implement in routine clinical care, it was reduced from its original 23 items (KCCQ-23) into a 12-item instrument (KCCQ-12), which includes the symptom frequency, physical limitations, social limitations, and quality of life domains and can also generate the clinical and overall summary scores with excellent concordance to the respective scores of the full instrument. Use of the 12 item questionnaire will reduce the response burden for patients.

#### 4 Perceived Stress Scale (PSS-10)

Stress and anxiety will be assessed subjectively using the PSS-10. Over a 10 item questionnaire it evaluates the degree to which an individual has perceived life as unpredictable, uncontrollable and overloading over the previous month. Participants fill out the questionnaire by rating the questions about their feelings and thoughts. The total score varies from 0 (no stress) to 40 (highest stress).

#### 5 State Trait Anxiety Inventory (STAI)

This is a psychological inventory consisting of 40 self-report items on a 4-point Likert scale. The STAI measures two types of anxiety – state anxiety and trait anxiety. Higher scores are positively correlated with higher levels of anxiety.

#### 6 Usability and Satisfaction analysis

In order to gather information on the acceptability and satisfaction with the cardio-oncology assessment and surveillance pathway, the opinions of patients will be obtained using a satisfaction survey. It is a Likert scale questionnaire evaluating seven dimensions of patient satisfaction including general satisfaction, technical quality, interpersonal manner, communication, financial aspects, time spent with doctor and accessibility and convenience. There will also be an opportunity for patients to share their perspectives freely with a free text option.

### Biosensors

Machine learning and stress monitoring is an area of translational research (19). Those patients identified in the baseline risk assessment as being at high risk of developing CTRCD will be offered wearable biosensors which will measure physiologic indicators of stress such as heart rate and blood pressure. The data from these wearable monitors (Huawei manufacture) will be correlated with chemical stress parameters (cortisol) and subjective assessments of stress in an effort to determine if the sensing technologies would be a feasible component of a future approach to monitoring stress which would facilitate early detection and intervention to prevent the progression of stress-aggravated cardiac conditions.

### Biomarkers

For patients undergoing neoadjuvant chemotherapy, biomarkers will be collected for biobanking and use in translational studies to identify cardiovascular and oncological biomarkers detectable in the tumor and/or circulation of this patient group, which may aid in the prognostication and personalization of therapeutic strategies on an individual patient basis. The schedule of biomarker sampling for these patients is outlined in the group protocols.

Biomarker evaluation will be via serum/plasma/whole blood collection, and will be drawn at the time of the patient’s clinic visit for cardiac risk assessment. This will include baseline measurement of Troponin I or Troponin T (based on local laboratory availability), and NT-proBNP or BNP (again based on laboratory availability). Samples will also be biobanked for future analysis of novel markers of CTRCD risk. The custodian of these samples will be the Cancer Biobank of Department of Surgery, Lambe Institute, University of Galway.

Patients deemed to be at high risk of CTRCD will also be asked to provide a sample of saliva for measurement of salivary cortisol at these time-points. This will then be correlated with other physiological parameters of stress measured using wearable sensors, and subjective measures of stress using questionnaires. Biomarker evaluation will also be requested in the event of an unexpected hospital admission, unplanned interruption in treatment or a cardiac event.

### Cardiac Imaging

TTE will be performed at baseline to inform risk stratification by providing quantitative assessment of biventricular size and function, presence of hypertrophy, regional wall motion abnormalities, diastolic function, atrial size, valvular heart disease, septal defects, pulmonary arterial pressure, assessment of the great vessels, and regarding any pericardial disease. The components of the baseline echocardiography study will be 2D and 3D image acquisition, global longitudinal strain, colour flow doppler, tissue doppler imaging, and pulsed/continuous wave doppler.

The feasibility of Cardiac Magnetic Resonance assessment of CTRCD will be sought in a subgroup of patients at high risk for cardiovascular events and those with non-interpretable or inadequate assessments on conventional ultrasound imaging. This is based on recommendation 4.1 (a) in the consensus document from European Society of Medical Oncology (20).

### Data Storage

All research staff working on this project, in addition to the Health Service Executive (HSE) investigators are trained in General Data Protection Regulation (GDPR) as per institutional requirement. Data collected will be specific to the study and not go beyond this.

All personal information provided for this research will be anonymized. The Clinical Research Facility Galway, Sligo cancer clinical research team, Mayo Cancer Nursing team, and the CORRIB Imaging Core Lab, in accordance with the data protection legislation and research legislation arising out of Good Clinical Practice for research, will process personal data such as name, surname, date and place of birth, address and the participants sensitive data, i.e. data relating to health and medical records, ethnic origin and lifestyle, only where necessary to achieve the objectives of the study.

A unique identifier will be assigned to the data. This data will not be processed in any way that will cause damage or distress to the participant. All personal data required and collected for this research is controlled by the institution where the patients are being cared for, and will be held on a hospital approved system to protect data. The files that link the participants name to the unique identifier will be stored in a secure location within the Clinical Research Facility in Galway. Staff from regulatory authorities or the Health Research Board Clinical Research Facility may also have access to the participants personal information for inspection and project oversight purposes. All parties who have access to the participant’s data are bound to maintain confidentiality in line with their employment contract and will have undertaken training in data protection law and practice. Data associated with this study may be used in the context of scientific research and publications or educational material but the participant’s identity will always be protected.

Data related to patient demographics, co-morbidities, medication and clinician record of actions/follow-up will be collected from the electronic patient record on an ongoing basis. Data relating to cardio-oncology services provided and used during the 24 month period will be extracted from patient records, as will data pertaining to compliance with recommended schedules of cardio-oncology surveillance during cancer treatment. Data on service utilisation outside of Galway University Hospital will be collected at the end of the study to include services used, frequency of use and personnel responsible for providing the service. This information will be collected by the research nurse. Blood samples will also be biobanked in the Cancer Biobank at University of Galway and may be utilised in studies analysing novel cardio-oncology biomarkers. These studies themselves will require project specific ethics to comply with general data protection regulations 2018.

### Potential risk

A risk assessment has been carried out, taking into account local policy. There is some level of low-level harm to be expected from taking blood samples e.g. pain, anxiety, bruising, infection, damage to local structures, excessive bleeding. Of note, patients will have study bloods taken at the same time as routine bloods and so there will be no requirement for additional phlebotomy.

Adverse events refer to any untoward event or medical occurrence that may not have a causal relationship with the treatment. This study is not evaluating medicinal interventions, therefore only the following adverse events only will be recorded:

- Cardiotoxic events/CTRCD (in accordance with 2023 ESC guideline definitions)
- Cancer re-occurrence
- Hospital admission or interruption to treatment

All staff including the Principal Investigator and study researchers have up to date training in Good Clinical Practice. A training log of completion is kept in the Clinical Research facility, with GDPR training compulsory for all involved staff. The study may also be audited internally and/or externally to ensure compliance. Any data breaches will follow local policy and standard operating procedure guidance on reporting and corrections In order to control the risk to each participants data, recording, transmission and storage of subjects’ trial-relevant data will be performed according to local secrecy obligations, as well as national and European requirements (European Union Directive 96/46 on data protection). The principal investigator will ensure that the patients anonymity is maintained. On documents submitted to the sponsor, subjects will not be identified by their names, but by their assigned identification number. It patient names are included on copies of documents submitted to the sponsor, the names will be obliterated and replaced with the assigned study patient numbers.

Patients are separately informed about data security in the patient information leaflets/informed consent form. The principal investigator will keep a separate log of patient identification numbers, names, addresses, telephone numbers and hospital numbers (if applicable) in the site file. Documents not for submission to the sponsor, such as signed informed consent forms, will be maintained in strict confidence by the principal investigator in the investigator site file (ISF). A screening failure log will be maintained for subjects who have consents to participate in the study but who, for whatever reason, are not eligible, withdrawn or decide to withdraw prior to taking part.

For electronic medical records the researcher will need their own individual access to the electronics systems. The staff members log in will record access. Personal data can be collected in two forms - one is collated as part of a trial and study and is called source data. This forms part of the patients medical records - as per typical clinical practices, a clinical personnel (e.g. a research nurse) enter notes into the patients record. This forms part of the medical notes and remains at the hospital site.

Other than the patients record - the only other location personnel data is kept is in the ISF. The original signed PIL and ICF for each study subject is filed in the ISF - this will have the patients’ signature and their name, address etc. Also to link a patient to their study code, a patient identification log is kept in the ISF. This contains the subjects name, chart number and date of birth, to link the patient to the trial specific code. To control access to this data, the primary investigator of the trial is responsible for the trial and the study. The ISF are retained in a secure office within the research/cancer department at Galway, Mayo or Sligo University Hospital, in a fireproof locked cabinet when not being worked on.

The primary investigator can delegate tasks to research personnel who are appropriately trained in the relevant area. Personnel data as before remains at the hospital site. Study files are archived under agreement with the institution. Outside of error correction, data is not erased or disclosed. Staff access to data is restricted to those delegated to work on a trial. The study will be monitored by the sponsor in addition the studies are open to internal and external audit and inspection to ensure compliance. Explicit consent will be obtained from the patients regarding access to her data.

Once the health research has been completed the pseudo-anonymised study data is either archived as required by legislation and local procedures by the sponsor and the ISF will be filed under agreement with Galway University Hospital. Study data will be maintained confidentially in a dedicated database in a secure location. This is primarily to facilitate subsequent follow-up of patients. It will be available for review by auditors or inspectors as required.

### Statistical Analysis

The PCORE Investigators have established collaboration with biostatisticians at the INSIGHT Science Foundation Ireland centre for data analytics within University of Galway for analysis of the multi-component dataset from UCARE. All analyses will be conducted and/or supervised by the collaborating statistician at Galway University under established quality systems and standard operating procedures, and in accordance with International Conference on Harmonization E9 Statistical Principles for Clinical Trials and E6 Good Clinical Practices.

Multivariate analysis will be performed for association between Health-related QOL measures and predictors in patients undergoing breast cancer treatment. We aim to prospectively enroll approximately 100 patients in this study. A sample size calculation has not been undertaken given that this study aims to evaluate the *feasibility* of introducing a cardio-oncology pathway for breast cancer patients. This follows established practice in pilot and feasibility studies as there are areas of uncertainty which this study will address to inform the sample size calculation of future randomized controlled trials to evaluate the efficacy of this pathway in early identification and management of CTRCD. Based on known clinical activity and number of breast cancer patients referred for chemotherapy annually, we believe 100 patients to be reasonable and achievable within the timeframe of this study.

### Declaration of results

Results will be made available through presentation at academic conferences and academic publication of results with colleagues in peer-reviewed journals. Results will also be communicated directly to key knowledge-users including the HSE and the National Cancer Control Programme (via local Medical Oncology Clinical Lead). This engagement will be critical to enable to healthcare services and policy sector make informed decisions or valuable changes to clinical practice, expenditure and/or systems development to support specialized Cardio-Oncology clinical pathways.

## Data Availability

All data is available upon request.

## Acknowledgements

Clinical Indemnity Insurance is provided by University Hospital Galway.

## Supplemental Materials

1 – Patient Information Leaflet/Informed Consent Form

2 – Group protocols

3 – General Practitioner Letter

4 – Letter to Medical Oncologist for high-risk patients

5 – Letter to Medical Oncologist for moderate-risk patients

6 – IPAQ questionnaire

7 – EORTC QLQ-BR45 questionnaire

8 – QLQ-C30 questionnaire

9 – KCCQ-12 questionnaire

10 – Perceived Stress Scale

11 – State Trait Anxiety Inventory

## Non-standard Abbreviations and Acronyms

CTRCD: Cancer Treatment Related Cardiac Dysfunction
ESC: European Society of Cardiology
QOL: Quality of life
ECG: Electrocardiogram
PIL: Patient information leaflet
ICF: Informed consent form
TTE: Transthoracic echocardiography
HFA-ICOS: Heart Failure Association – International Cardio-Oncology Society
EORTC: The European Organisation for Research and Treatment of Cancer
IPAQ-SF: International Physical Activity Questionnaire Short Form
KCCQ: Kansas City Cardiomyopathy Questionnaire
PSS-10: Perceived Stress Scale
STAI: State Trait Anxiety Inventory
HSE: Health Service Executive
GDPR: General Data Protection Regulation
ISF: Investigator site file

